# Sports injuries and illnesses during the Beijing 2022 Winter Olympic Games in both athletes and nonathletes in the Zhangjiakou Competition Zone

**DOI:** 10.1101/2022.12.09.22283286

**Authors:** Zhen-long Liu, Can-can Du, Jian-li Gao, Yue-yang Hou, Ya-jie Wang, Yu-han Zhang, Yan-fang Jiang, Yu-ping Yang

## Abstract

**Objectives:** To describe the incidences of injuries and illnesses sustained during the XXIV Winter Olympic Games, hosted by Beijing in 2022.

**Methods:** We recorded the daily number of injuries and illnesses in athletes and nonathletes in the polyclinic and medical venues documented by the Beijing 2022 medical staff.

**Results:** In total, 35,537 athletes and nonathletes were evaluated for injuries and illnesses. The Beijing 2022 medical staff reported 725 injuries and 1119 illnesses comprising 24 injuries and 31 illnesses per 1000 athletes and nonathletes during the Winter Olympic Games. Altogether, 2.4% of the athletes and nonathletes incurred at least one injury and 3.1% had at least one illness. The injury incidence was highest in athletes (14.4%), team officials (8.8%), staff (2.1%), and volunteers (2.1%), and lowest in national and international technical officials (1.5%) and stakeholders (0.3%). The highest incidences of illness were recorded in team officials (7.5%), volunteers (4.8%), and athletes (4.6%). Seventeen percent of the illnesses and injuries affected the oral structures, 15.4% affected the gastrointestinal system, and 10.5% affected the respiratory system.

**Conclusion:** The injury morbidity rate of athletes was 14.4%, and the illness morbidity rate was 4.6%. For nonathletes, the injury morbidity rate was 2.0%, and the illness morbidity rate was 3.1%. The incidences of injuries and illnesses varied substantially among the different categories of participants. Analyses of the injury mechanisms and illness causes in athletes and nonathletes during the Winter Olympic Games are essential to devise better injury and illness prevention strategies.

**Summary Box:**

- Injury and illness rates have been reported for previous Winter Olympic Games but not for the Beijing 2022 Winter Olympic Games.
- Previous studies of Winter Olympic Games focused on injuries in athletes, only. In this study, we evaluated injury and illness rates in both athletes and nonathletes.
- With the results of this study, we hoped to provide a reference for the formulation of diagnostic and treatment strategies for injuries and illnesses for athletes and nonathletes participating in the Winter Olympic Games.

## INTRODUCTION

The Beijing 2022 Winter Olympic Games was a large world sporting event with more than 35,000 participating athletes and nonathletes from more than 90 countries. There were three competition zones for the Beijing 2022 Winter Olympic Games. As the main competition venue, the Zhangjiakou Competition Zone was the location of two major disciplines and six events, including freestyle skiing, biathlon, ski jumping, and cross-country skiing. Fifty-one gold medals were won at this venue, which was the largest number of gold medals in the three competition zones. The competition required extensive manpower to address needs related to weather, medical care, and transport and catering, owing to the extremely low temperature and high requirement for venue security. The Beijing 2022 Winter Olympic Games were held successfully because 35,537 security personnel (nonathletes), played a significant role. There was a high number of athletes and nonathletes at these games, and the environment was challenging. Consequently, injuries and diseases were inevitable. Additionally, coronavirus disease-2019 (COVID-19) was still spreading around the world at the time of the games. To reduce the infection risk to athletes and nonathletes, closed-loop management was implemented, which increased the requirement for medical security during the competition. The Zhangjiakou Winter Olympic Village Polyclinic and Peking University Third Hospital-Chongli provided medical security for athletes and nonathletes. The Winter Olympic Village Polyclinic mainly diagnosed and treated patients with mild injuries and illnesses, while those with serious injuries and illnesses were transferred to Peking University Third Hospital-Chongli. The medical security system provided reasonable and efficient diagnosis and treatment for athletes and nonathletes.

The International Olympic Committee (IOC) increasingly emphasizes the protection of athletes’ health and the prevention of injuries. At the Beijing 2008 Olympic Games, the IOC commissioned the first major IOC injury surveillance system^1, 2^, which was subsequently expanded for Vancouver 2010 to also include illnesses^3^. The surveillance system was further developed for the London 2012^4^, Sochi 2014^5^, Rio 2016^6^, and PyeongChang 2018^7^ games. However, there are few reports of injuries and illnesses in Olympic nonathletes for previous Olympic Games. Therefore, we performed a study of injuries and illnesses in both athletes and nonathletes who were evaluated in the Zhangjiakou Competition Zone’s medical institutions. We analyzed the epidemiology in terms of the injury types, locations, and causes, and the affected systems, symptoms, and causes, for illnesses. Our aim was to describe the incidence and characteristics of the sports injuries and illnesses that occurred during the Beijing 2022 Winter Olympic Games^8^.

## METHODS

We used the IOC injury and illness surveillance system for multisport events in this observational study^1^. All patients’ medical histories were recorded using the electronic medical record system. We retrieved information for athletes and nonathletes treated for injuries and illnesses in the Winter Olympic Village Polyclinic and Peking University Third Hospital-Chongli in the Zhangjiakou Competition Zone of the Beijing 2022 Winter Olympic Games^9^.

### Implementation

#### Definition of injury and illness

We defined injuries and illnesses as new (pre-existing, not fully rehabilitated conditions were not recorded) or recurring (patients having returned to full participation after a previous condition) musculoskeletal symptoms or concussions (injuries) or illnesses incurred during security work, competition, or training during the Beijing 2022 Winter Olympic Games (4–20 February 2022)^10^. We obtained data for people receiving medical attention, regardless of the consequences of absence from competition or training^1^. In cases where a single incident caused multiple injury types or affected multiple body parts, we recorded all diagnoses, as determined by our research team^4^.

### Injury and illness case report form

Our injury and illness case report form followed the template of that used in the Vancouver 2010, London 2012, Sochi 2014, Rio 2016, and PyeongChang 2018 Olympic Games^3-7^. With respect to injuries and illnesses, we recorded the following data: name, age, personnel categories, sports and events, vital signs, medical history, diagnosis, physical examination findings, auxiliary examinations and findings, treatments, date and time, injury locations, types, and causes, affected systems, main symptoms, and causes.

### Equity, diversity, and inclusion statement

We analyzed data for all patients who visited the Winter Olympic Village Polyclinic and Peking University Third Hospital-Chongli during the Beijing 2022 Winter Olympic Games. We focused on the incidence of injuries and illnesses according to the location and system involved. However, we did not compare the morbidity by sex. We did not purposefully recruit people from marginalized communities. Our author team comprised three women and five men, all of whom were from China. The authors’ discipline was sports medicine. Yu-ping Yang is medical expert of sports medicine working for Beijing 2022 Winter Olympic Games. As sports medicine experts, Zhen-long Liu and Yan-fang Jiang participated in the medical work of the Beijing 2022 Winter Olympic Games. The remaining researchers had extensive research experience. Our analysis explored the morbidity of sports injuries and illnesses of athletes and nonathletes. However, we did not examine the effects of ethnicity. We discussed the influence of the profession of the athletes and nonathletes on our findings in the discussion.

### Confidentiality

We retrieved information for athletes and nonathletes who were treated for injuries and illnesses in the medical institutions. These data were collected through an electronic medical record system. We treated all information with strict confidence, and anonymized our medical database at the end of the Games.

### Data analysis

All data were analyzed statistically using SPSS (SPSS for Windows, version 25.0; IBM Corp., Armonk, IL, USA). Enumeration data were presented as the number of cases and percentage for baseline information, age distribution, number and proportions of injury types, locations, and causes, and number and proportions of illness-affected systems, symptoms and causes, rapid diagnostic testing, and medical treatments. Measurement data, such as age distribution, were presented as means and standard deviations.

## RESULTS

The total number of athletes in the Zhangjiakou Competition Zone was 1222. The total number of nonathletes in the closed loop was 35,537. Among the nonathletes, 20,911 were staff, 2165 were national technical officials (NTOs) and international technical officials (ITOs), 4500 were volunteers, 1127 were team officials, and 5612 were stakeholders. We recorded 1119 illnesses and 725 injuries documented in the polyclinic and medical venues. The Zhangjiakou Winter Olympic Village Polyclinic evaluated 957 athletes and nonathletes with 611 illnesses and 346 injuries. Peking University Third Hospital-Chongli evaluated 887 athletes and nonathletes with 508 illnesses and 379 injuries.

### Baseline information

The baseline information of the different personnel categories is shown in Table 1. The baseline information comprised age, sex, and the injury and illness distributions of the different categories of personnel. The total number of doctor visits was 1844 and these comprised visits by athletes (n=198, 10.7%), staff (n=1050, 56.9%), NTOs and ITOs (n=56, 3.0%), volunteers (n=294, 15.9%), team officials (n=177, 9.6%), and stakeholders (n=69, 3.7%). The injured/ill athletes comprised 90 women, representing 13.2% of all women in the study. The 108 male athletes who were evaluated for injury or illness represented 9.3% of all men in the study. The injured/ill staff comprised 356 women, representing 52.2% of all women in the study. The 694 injured/ill men represented 59.7% of all men in the study.

**Table 1.** Baseline information of different personnel category

| Personnel category | Age Mean±SD | Female n*=682 (%) | Male n*=1162 (%) | Injury n*=725 (%) | Illness n*=1119 (%) | Total n*=1844 (%) |
| --- | --- | --- | --- | --- | --- | --- |
| Athletes | 25.8±6.1 | 90(13.2) | 108(9.3) | 133(18.3) | 65(5.8) | 198(10.7) |
| Staff | 34.5±11.6 | 356(52.2) | 694(59.7) | 384(53.0) | 666(59.5) | 1050(56.9) |
| NTO and ITO | 39.0±12.4 | 21(3.1) | 35(3.0) | 27(3.7) | 29(2.6) | 56(3.0) |
| Volunteers | 32.0±9.1 | 152(22.3) | 142(12.2) | 73(10.1) | 221(19.7) | 294(15.9) |
| Team officials | 46.0±13.3 | 44(6.5) | 133(11.4) | 93(12.8) | 84(7.5) | 177(9.6) |
| Stakeholder | 33.9±9.4 | 19(2.8) | 50(4.3) | 15(2.1) | 54(4.8) | 69(3.7) |
| Total | 34.4±11.8 | 682(37.0) | 1162(63.0) | 725(39.3) | 1119(60.7) | 1844 |

\*means frequency of doctor visits

Table 2 shows the results of the analysis of the age distribution of the different categories of personnel. The percentages of the different categories in each age group are shown in parentheses. Athletes suffered more injuries than illnesses, and all other personnel categories developed more illnesses than injuries. It was obvious that athletes were generally younger than other people seen at the medical facilities, with 193 (97.5%) under the age of 40 years. In contrast to the athletes, the team officials were generally older, with 119 (67.2%) over the age of 40 years.

**Table 2.**
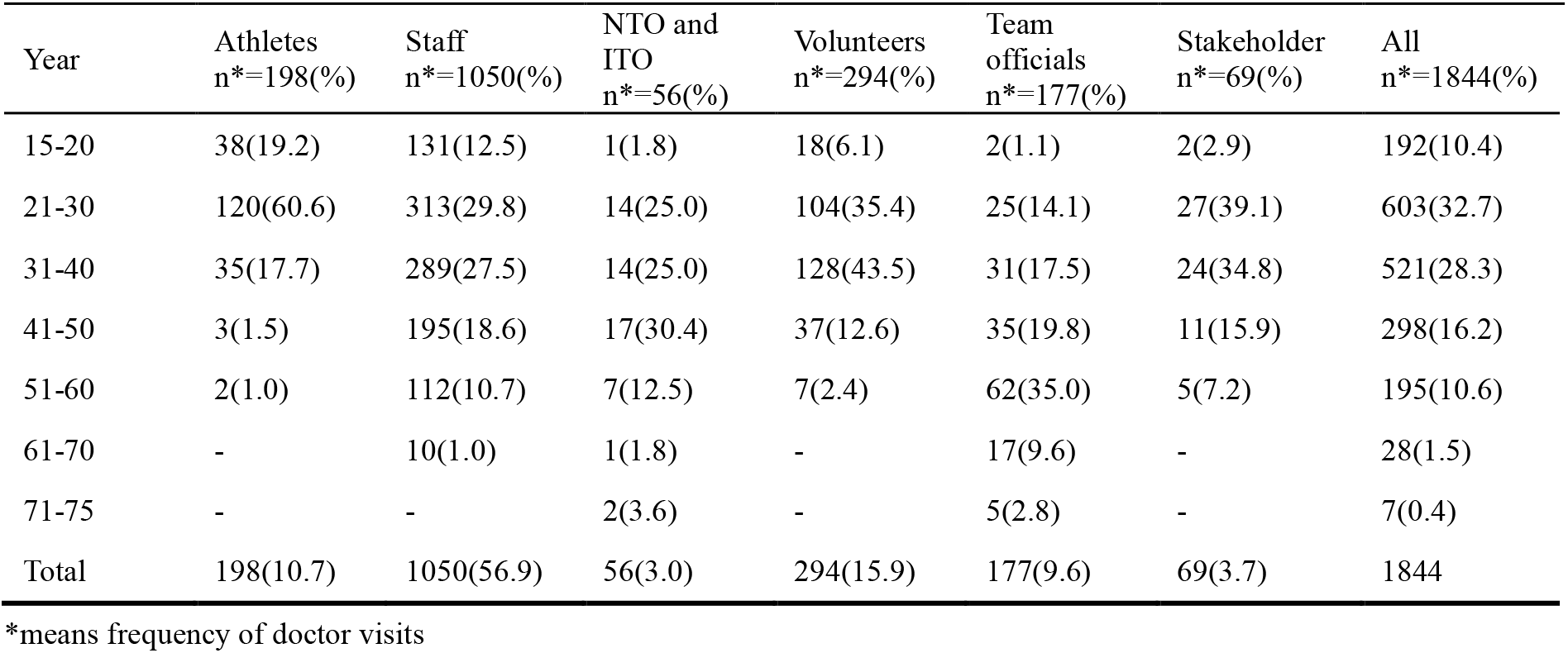
Age distribution of different personnel category

| Year | Athletes n*=198(%) | Staff n*=1050(%) | NTO and ITO n*=56(%) | Volunteers n*=294(%) | Team officials n*=177(%) | Stakeholder n*=69(%) | All n*=1844(%) |
| --- | --- | --- | --- | --- | --- | --- | --- |
| 15-20 | 38(19.2) | 131(12.5) | 1(1.8) | 18(6.1) | 2(1.1) | 2(2.9) | 192(10.4) |
| 21-30 | 120(60.6) | 313(29.8) | 14(25.0) | 104(35.4) | 25(14.1) | 27(39.1) | 603(32.7) |
| 31-40 | 35(17.7) | 289(27.5) | 14(25.0) | 128(43.5) | 31(17.5) | 24(34.8) | 521(28.3) |
| 41-50 | 3(1.5) | 195(18.6) | 17(30.4) | 37(12.6) | 35(19.8) | 11(15.9) | 298(16.2) |
| 51-60 | 2(1.0) | 112(10.7) | 7(12.5) | 7(2.4) | 62(35.0) | 5(7.2) | 195(10.6) |
| 61-70 | - | 10(1.0) | 1(1.8) | - | 17(9.6) | - | 28(1.5) |
| 71-75 | - | - | 2(3.6) | - | 5(2.8) | - | 7(0.4) |
| Total | 198(10.7) | 1050(56.9) | 56(3.0) | 294(15.9) | 177(9.6) | 69(3.7) | 1844 |

\*means frequency of doctor visits

### Incidence of injuries

Among the 35,537 registered athletes and nonathletes, 864 injuries were reported, resulting in an injury rate of 24.3 injuries per 1000 athletes and nonathletes. On average, 2.4% of the athletes and nonathletes sustained at least one injury (n=864). Athletes experienced the highest incidence of injuries, resulting in an injury rate of 144 injuries per 1000 registered athletes. Similarly, there were 88 injuries per 1000 team officials, 21 injuries per 1000 for both staff and volunteers, 15 injuries per 1000 for NTOs and ITOs, combined, and 3 injuries per 1000 stakeholders (Table 3).

**Table 3.**
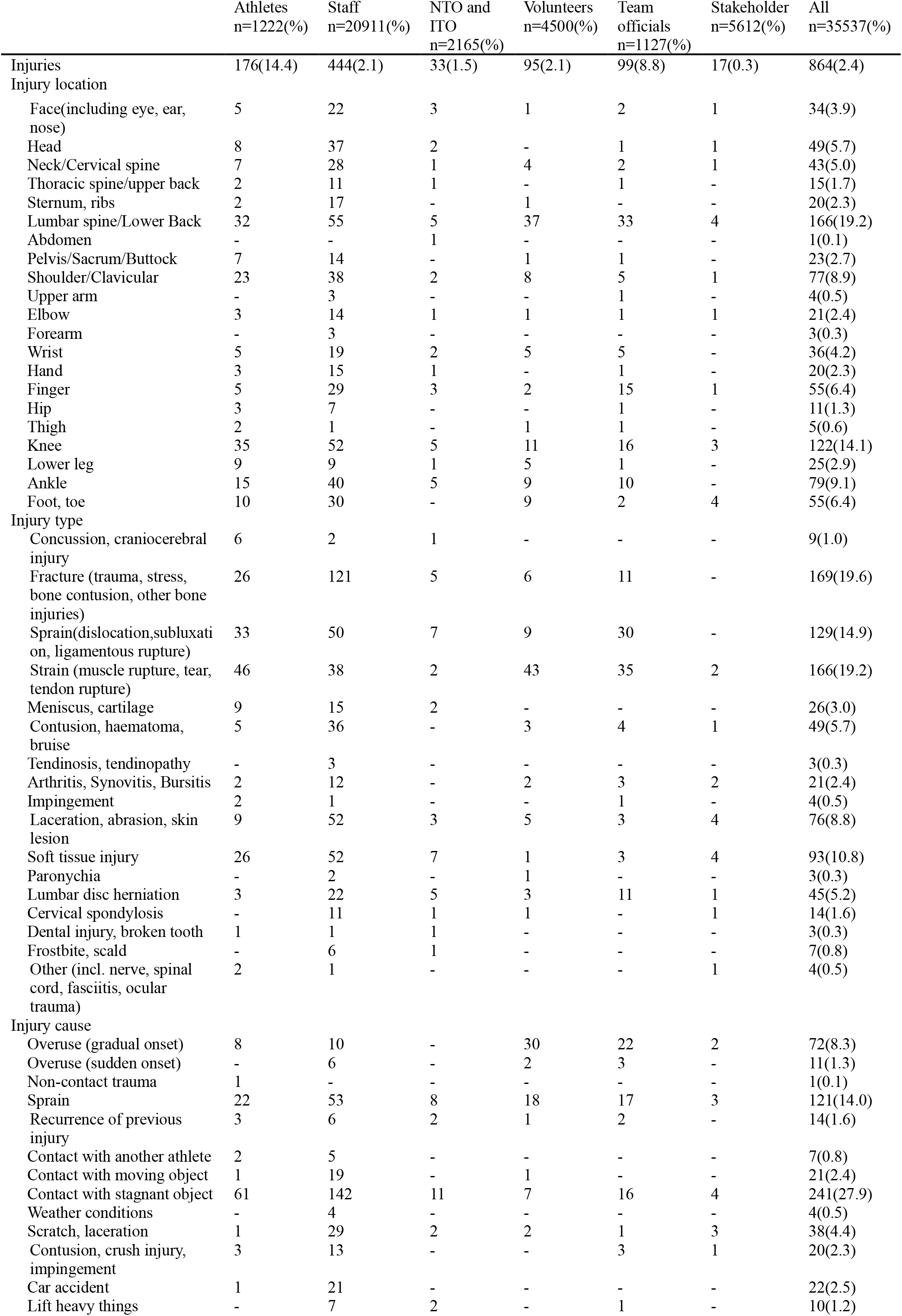
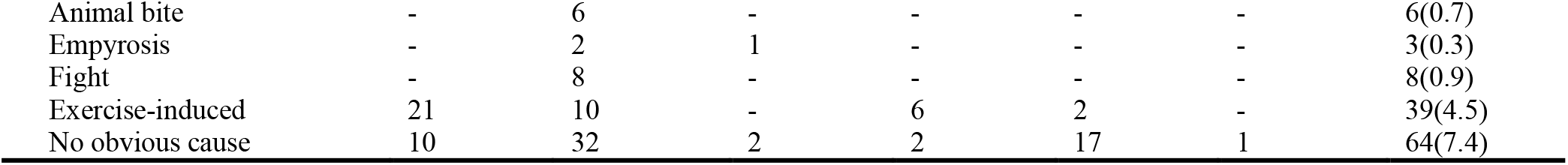
Number (n) and proportions (%) of injury types, locations and causes

|  | Athletes<br>n=1222(%) | Staff<br>n=20911(%) | NTO and<br>ITO<br>n=2165(%) | Volunteers<br>n=4500(%) | Team<br>officials<br>n=1127(%) | Stakeholder<br>n=5612(%) | All<br>n=35537(%) |
| --- | --- | --- | --- | --- | --- | --- | --- |
| Injuries | 176(14.4) | 444(2.1) | 33(1.5) | 95(2.1) | 99(8.8) | 17(0.3) | 864(2.4) |
| Injury location |  |  |  |  |  |  |  |
| Face(including eye, ear,<br>nose) | 5 | 22 | 3 | 1 | 2 | 1 | 34(3.9) |
| Head | 8 | 37 | 2 | - | 1 | 1 | 49(5.7) |
| Neck/Cervical spine | 7 | 28 | 1 | 4 | 2 | 1 | 43(5.0) |
| Thoracic spine/upper back | 2 | 11 | 1 | - | 1 | - | 15(1.7) |
| Sternum, ribs | 2 | 17 | - | 1 | - | - | 20(2.3) |
| Lumbar spine/Lower Back | 32 | 55 | 5 | 37 | 33 | 4 | 166(19.2) |
| Abdomen | - | - | 1 | - | - | - | 1(0.1) |
| Pelvis/Sacrum/Buttock | 7 | 14 | - | 1 | 1 | - | 23(2.7) |
| Shoulder/Clavicular | 23 | 38 | 2 | 8 | 5 | 1 | 77(8.9) |
| Upper arm | - | 3 | - | - | 1 | - | 4(0.5) |
| Elbow | 3 | 14 | 1 | 1 | 1 | 1 | 21(2.4) |
| Forearm | - | 3 | - | - | - | - | 3(0.3) |
| Wrist | 5 | 19 | 2 | 5 | 5 | - | 36(4.2) |
| Hand | 3 | 15 | 1 | - | 1 | - | 20(2.3) |
| Finger | 5 | 29 | 3 | 2 | 15 | 1 | 55(6.4) |
| Hip | 3 | 7 | - | - | 1 | - | 11(1.3) |
| Thigh | 2 | 1 | - | 1 | 1 | - | 5(0.6) |
| Knee | 35 | 52 | 5 | 11 | 16 | 3 | 122(14.1) |
| Lower leg | 9 | 9 | 1 | 5 | 1 | - | 25(2.9) |
| Ankle | 15 | 40 | 5 | 9 | 10 | - | 79(9.1) |
| Foot, toe | 10 | 30 | - | 9 | 2 | 4 | 55(6.4) |
| Injury type |  |  |  |  |  |  |  |
| Concussion, craniocerebral<br>injury | 6 | 2 | 1 | - | - | - | 9(1.0) |
| Fracture (trauma, stress,<br>bone contusion, other bone<br>injuries) | 26 | 121 | 5 | 6 | 11 | - | 169(19.6) |
| Sprain(dislocation,subluxati<br>on, ligamentous rupture) | 33 | 50 | 7 | 9 | 30 | - | 129(14.9) |
| Strain (muscle rupture, tear,<br>tendon rupture) | 46 | 38 | 2 | 43 | 35 | 2 | 166(19.2) |
| Meniscus, cartilage | 9 | 15 | 2 | - | - | - | 26(3.0) |
| Contusion, haematoma,<br>bruise | 5 | 36 | - | 3 | 4 | 1 | 49(5.7) |
| Tendinosis, tendinopathy | - | 3 | - | - | - | - | 3(0.3) |
| Arthritis, Synovitis, Bursitis | 2 | 12 | - | 2 | 3 | 2 | 21(2.4) |
| Impingement | 2 | 1 | - | - | 1 | - | 4(0.5) |
| Laceration, abrasion, skin<br>lesion | 9 | 52 | 3 | 5 | 3 | 4 | 76(8.8) |
| Soft tissue injury | 26 | 52 | 7 | 1 | 3 | 4 | 93(10.8) |
| Paronychia | - | 2 | - | 1 | - | - | 3(0.3) |
| Lumbar disc herniation | 3 | 22 | 5 | 3 | 11 | 1 | 45(5.2) |
| Cervical spondylosis | - | 11 | 1 | 1 | - | 1 | 14(1.6) |
| Dental injury, broken tooth | 1 | 1 | 1 | - | - | - | 3(0.3) |
| Frostbite, scald | - | 6 | 1 | - | - | - | 7(0.8) |
| Other (incl. nerve, spinal<br>cord, fasciitis, ocular<br>trauma) | 2 | 1 | - | - | - | 1 | 4(0.5) |
| Injury cause |  |  |  |  |  |  |  |
| Overuse (gradual onset) | 8 | 10 | - | 30 | 22 | 2 | 72(8.3) |
| Overuse (sudden onset) | - | 6 | - | 2 | 3 | - | 11(1.3) |
| Non-contact trauma | 1 | - | - | - | - | - | 1(0.1) |
| Sprain | 22 | 53 | 8 | 18 | 17 | 3 | 121(14.0) |
| Recurrence of previous<br>injury | 3 | 6 | 2 | 1 | 2 | - | 14(1.6) |
| Contact with another athlete | 2 | 5 | - | - | - | - | 7(0.8) |
| Contact with moving object | 1 | 19 | - | 1 | - | - | 21(2.4) |
| Contact with stagnant object | 61 | 142 | 11 | 7 | 16 | 4 | 241(27.9) |
| Weather conditions | - | 4 | - | - | - | - | 4(0.5) |
| Scratch, laceration | 1 | 29 | 2 | 2 | 1 | 3 | 38(4.4) |
| Contusion, crush injury,<br>impingement | 3 | 13 | - | - | 3 | 1 | 20(2.3) |
| Car accident | 1 | 21 | - | - | - | - | 22(2.5) |
| Lift heavy things | - | 7 | 2 | - | 1 | - | 10(1.2) |
| Animal bite | - | 6 | - | - | - | - | 6(0.7) |
| Empyrosis | - | 2 | 1 | - | - | - | 3(0.3) |
| Fight | - | 8 | - | - | - | - | 8(0.9) |
| Exercise-induced | 21 | 10 | - | 6 | 2 | - | 39(4.5) |
| No obvious cause | 10 | 32 | 2 | 2 | 17 | 1 | 64(7.4) |

### Injury location and type

For injuries to both athletes and nonathletes, the lumbar spine and lower back (n=166, 19.2%) were the most prominent injury locations, followed by the knee (n=122, 14.1%), ankle (n=79, 9.1%), and shoulder and clavicle (n=77, 8.9%) (Table 3). The three most common injury locations in athletes were the knee (n=35, 19.9%), lumbar spine and lower back (n=32, 18.2%), and shoulder and clavicle (n=23, 13.1%). For the staff, the most common injury locations were the lumbar spine and lower back (n=55, 12.4%), knee (n=52, 11.7%), and ankle (n=40, 9.0%).

Fracture (trauma, stress, bone contusion, other bone injuries; n=169, 19.6%), strain (muscle rupture, tear, tendon rupture; n=166, 19.2%), sprain (dislocation, subluxation, ligamentous rupture; n=129, 14.9%), and soft tissue injury (n=93, 10.8%) were the most common injury types overall (Table 3).

### Injury cause

Eighteen mechanisms of injury were reported, and the three most common were contact with a stationary object (n=241, 27.9%), sprain (n=121, 14.0%), and overuse (gradual onset) (n=72, 8.3%) (Table 3). Details on the other mechanisms of injury are shown in Table 3.

### Incidence and distribution of illnesses

Among 1844 of the 35,537 athletes and nonathletes (5.2%), 1103 illnesses were reported, resulting in an incidence of 31 illnesses per 1000 athletes and nonathletes. The numbers (n) and proportions (%) of illnesses in each group were analyzed individually. The three most common groups for whom illnesses were reported were team officials (n=84, 7.5%), volunteers (n=218, 4.8%), and athletes (n=56, 4.6%) (Table 4).

**Table 4.** Number (n) and proportions (%) of illness affected systems, symptoms and causes

|  |  |  |  |  |  |  |  |
| --- | --- | --- | --- | --- | --- | --- | --- |
|  | Athletes<br>n=1222(%) | Staff<br>n=20911(%) | NTO and<br>ITO<br>n=2165(%) | Volunteers<br>n=4500(%) | Team<br>officials<br>n=1127(%) | Stakeholder<br>n=5612(%) | All<br>n=35537(%) |
| Illness | 56(4.6) | 665(3.2) | 27(1.2) | 218(4.8) | 84(7.5) | 53(0.9) | 1103(3.1) |
| Illness affected system |  |  |  |  |  |  |  |
| Gastrointestinal | 9 | 103 | 7 | 32 | 9 | 10 | 170(15.4) |
| Respiratory | 5 | 64 | 3 | 37 | 4 | 3 | 116(10.5) |
| Allergic, immunological | - | 1 | 1 | - | - | - | 2(0.2) |
| Metabolic, endocrinological | 1 | 28 | - | 5 | 3 | - | 37(3.4) |
| Hematologic | 2 | 4 | 1 | - | 1 | - | 8(0.7) |
| Dermatologic | 2 | 78 | - | 26 | 7 | 2 | 115(10.4) |
| Urogenital | 7 | 38 | 1 | 1 | 1 | - | 48(4.4) |
| Gynaecological | 3 | 18 | - | 5 | 3 | 4 | 33(3.0) |
| Cardiovascular | 1 | 88 | 6 | 9 | 3 | 8 | 115(10.4) |
| Vasculature | - | 2 | 2 | 1 | 1 | - | 6(0.5) |
| Neurological | 2 | 42 | 2 | 30 | 1 | 3 | 80(7.3) |
| Musculoskeletal | 1 | 15 | 1 | - | - | - | 17(1.5) |
| Stomatology | 12 | 78 | 1 | 46 | 39 | 12 | 188(17.0) |
| Eye | 5 | 53 | 1 | 11 | 7 | 8 | 85(7.7) |
| Otolaryngology | - | 48 | 1 | 15 | 3 | 3 | 70(6.3) |
| Other (intoxication,<br>thermoregulatory system) | 6 | 5 | - | - | 2 | - | 13(1.2) |
| Illness symptom |  |  |  |  |  |  |  |
| Fever, chill | 2 | 10 | - | 1 | 1 | 2 | 16(1.5) |
| Abdominal pain | 4 | 83 | 4 | 11 | 5 | 7 | 114(10.3) |
| Abdominal distension | 2 | 8 | - | 4 | - | 1 | 15(1.4) |
| Anorexia | - | 5 | - | 2 | - | - | 7(0.6) |
| Nausea | 1 | 55 | - | 8 | 1 | 1 | 66(6.0) |
| Vomiting | 2 | 47 | 1 | 5 | 1 | - | 56(5.1) |
| Diarrhoea | 1 | 20 | - | 2 | 4 | 4 | 31(2.8) |
| Constipation | 1 | 4 | - | 6 | - | - | 11(1.0) |
| Belching | 1 | 9 | - | 7 | 1 | 1 | 19(1.7) |
| Dyspnoea | - | 23 | 4 | 2 | - | - | 29(2.6) |
| Cough | 2 | 28 | - | 14 | 2 | 2 | 48(4.4) |
| Expectorine | - | 8 | - | 1 | 1 | 1 | 11(1.0) |
| Pharyngalgia | 3 | 25 | - | 17 | 4 | 1 | 50(4.5) |
| Nasal obstruction | 1 | 5 | - | 19 | 2 | - | 27(2.4) |
| Runny nose, sneeze | - | 7 | - | 20 | 2 | - | 29(2.6) |
| Chest pain | - | 26 | 2 | 5 | - | 1 | 34(3.1) |
| Chest stuffiness | - | 23 | 5 | 1 | - | 1 | 30(2.7) |
| Dizziness, syncope | 2 | 64 | 3 | 6 | - | - | 75(6.8) |
| Headache | - | 17 | 1 | 6 | 2 | 2 | 28(2.5) |
| Backache | - | 20 | 3 | - | - | - | 23(2.1) |
| Palpitation | - | 31 | 1 | 6 | 1 | - | 39(3.5) |
| Hypertension | - | 10 | - | - | 2 | 7 | 19(1.7) |
| Rash, pruritus | 3 | 70 | - | 25 | 6 | 5 | 109(9.9) |
| Red and swollen | 1 | 22 | - | 5 | 2 | 1 | 31(2.8) |
| Pain | 1 | 19 | - | 2 | - | - | 22(2.0) |
| Subcutaneous mass | - | 9 | 2 | 2 | - | - | 13(1.2) |
| Colporrhagia | 2 | 10 | - | 1 | 2 | - | 15(1.4) |
| Menelipsis, dysmenorrhea | - | 5 | - | 4 | - | 1 | 10(0.9) |
| Anal pain, hematochezia | 2 | 6 | 2 | 1 | 1 | - | 12(1.1) |
| Perianal abscess, haemorrhoid | 1 | 10 | - | 3 | - | - | 14(1.3) |
| Epistaxis | - | 9 | - | 2 | - | 2 | 13(1.2) |
| Hematuresis | - | 6 | - | - | - | - | 6(0.5) |
| Urinary urgency and frequency, odynuria, dysuresia | 5 | 6 | - | - | - | - | 11(1.0) |
| Blurred vision | 3 | 11 | - | - | 4 | 2 | 20(1.8) |
| Eye pain | - | 21 | - | 3 | - | 1 | 25(2.3) |
| Eyes itch | - | 4 | - | 2 | 1 | - | 7(0.6) |
| Lacrimation | - | 4 | - | - | 1 | - | 5(0.5) |
| Foreign body sensation | - | 16 | 1 | - | - | - | 17(1.5) |
| Conjunctival congestion | 2 | 5 | 2 | 2 | - | 2 | 13(1.2) |
| Dry eyes | - | 6 | - | 2 | 3 | 3 | 14(1.3) |
| Earache | - | 9 | - | 4 | - | 1 | 14(1.3) |
| Ears itch | - | 5 | - | - | - | - | 5(0.5) |
| Hearing loss | - | 5 | - | - | - | - | 5(0.5) |
| Tinnitus | - | 9 | - | 1 | 1 | - | 11(1.0) |
| Toothache | 7 | 70 | 1 | 42 | 36 | 11 | 167(15.1) |
| Oral ulcer | - | 1 | - | 3 | - | 1 | 5(0.5) |
| Neuritis, numb | - | 10 | 2 | - | - | - | 12(1.1) |
| Musculoskeletal pain | - | 21 | - | 3 | 1 | - | 25(2.3) |
| Weak, hidrosis | 2 | 22 | - | 2 | 1 | - | 27(2.4) |
| Insomnia | 1 | 10 | 1 | 24 | - | 1 | 37(3.4) |
| Hyperglycemia | - | 4 | - | 2 | - | - | 6(0.5) |
| No obvious symptoms | 16 | 20 | 1 | 4 | 6 | - | 47(4.3) |
| Other (including emaciation, jaundice, twitch, alcoholism, venous thrombosis, edema, morning stiffness, cardiopulmonary arrest) | - | 13 | 3 | - | 1 | - | 17(1.5) |
| **Illness cause** |  |  |  |  |  |  |  |
| Infection | 34 | 562 | 26 | 186 | 73 | 43 | 924(83.8) |
| Exercise-induced | 2 | 7 | - | - | - | - | 9(0.8) |
| Pre-existing | 1 | 15 | 2 | 3 | 2 | 4 | 27(2.4) |
| Overworked | - | 3 | - | - | - | - | 3(0.3) |
| Ultraviolet ray | - | 7 | - | - | - | - | 7(0.6) |
| Alcohol drinking | - | 5 | 1 | - | - | - | 6(0.5) |
| Menstrual period, abortion | - | 8 | - | - | - | 1 | 9(0.8) |
| No obvious cause | 20 | 66 | - | 32 | 9 | 5 | 132(12.0) |
| Other (including food, drug, cosmetics, foreign body) | - | 12 | - | - | - | - | 12(1.1) |

### Affected system, main symptoms, and causes of illness

A total of 188 illnesses (17.0%) affected the oral structures. The second and third most frequently affected systems were the digestive system (n=170, 15.4%) and the respiratory system (n=116, 10.5%), respectively. The fourth and fifth most frequently affected systems were the cardiovascular system (n=115, 10.4%) and dermatological system (n=115, 10.4%), respectively (Table 4).

The most common symptom was toothache (n=167, 15.1%). The second and third most common symptoms were abdominal pain (n=114, 10.3%) and rash and pruritus (n=109, 9.9%), respectively. The fourth and fifth most common symptoms were dizziness and syncope (n=75, 6.8%) and nausea (n=66, 6.0%), respectively (Table 4).

Infection was the most common cause of illness (n=924, 83.8%). The second and third causes were no obvious cause (n=132, 12.0%) and pre-existing illness (n=27, 2.4%), respectively. The distribution of affected systems, main symptoms, and causes of illness per participant/personnel category are presented in Table 4.

### Auxiliary examinations and laboratory tests

A total of 1401 auxiliary examinations and laboratory tests were performed in the polyclinic and medical venues. The five most common auxiliary examinations were digital radiography (n=296, 21.1%), computed tomography (n=240, 17.1%), blood routine examination (n=210, 15.0%), blood biochemistry (n=200, 14.3%), and ultrasonography (n=149, 10.6%) (Table 5).

**Table 5.** Number (n) and proportions (%) of auxiliary examinations and lab tests

|  |  |  |  |
| --- | --- | --- | --- |
| Auxiliary examination and lab tests | Number<br>n=1401(%) | Auxiliary examination and lab tests | Number<br>n=1401(%) |
| Digital radiography | 296(21.1) | Routine urine test | 37(2.6) |
| Computed tomography | 240(17.1) | Stool routine | 4(0.3) |
| Magnetic resonance | 99(7.1) | Blood biochemistry | 200(14.3) |
| Ultrasound | 149(10.6) | Arterial blood gas analysis | 6(0.4) |
| Gastrointestinal endoscope | 2(0.1) | Immunologic test | 1(0.1) |
| Laryngoscope | 3(0.2) | Optometry | 4(0.3) |
| Otoscope | 1(0.1) | Fundus photography | 3(0.2) |
| Electrocardiograph | 144(10.3) | Irrigation of lacrimal passage | 2(0.1) |
| Blood routine examination | 210(15.0) | Total | 1401 |

### Medical treatments

Medical treatments comprised dispensing over-the-counter and prescription medications, providing manual therapy, taping and bracing, and prescribing therapeutic physical therapy modalities. The three most common medications were antimicrobials (n=149, 21.32%), cardiovascular medications (n=67, 9.59%), and antipyretic/analgesics (n=61, 8.73%) (Table 6).

**Table 6.** Number (n) and proportions (%) of medical treatments

| **Table 6** Number (n) and proportions (%) of medical treatments |  |  |  |
| --- | --- | --- | --- |
| Prescription medications | Number<br>n=699(%) | Prescription medications | Number<br>n=699(%) |
| Immunotherapy | 42(6.01) | Gastrointestinal medications | 55(7.87) |
| Non-steroidal anti-inflammatories | 28(4.01) | Respiratory drugs | 16(2.29) |
| Glucocorticoid | 48(6.87) | Endocrine system medications | 19(2.72) |
| Antipyretic/analgesics | 61(8.73) | Ear/nose/throat drugs | 8(1.14) |
| Antimicrobials | 149(21.32) | Eye medications | 50(7.15) |
| Antiviral agent | 31(4.43) | Skin medications | 10(1.43) |
| Antihistamines | 41(5.87) | Nervous system medications | 29(4.15) |
| Cardiovascular medications | 67(9.59) | Musculoskeletal medications | 39(5.58) |
| Reproductive system drugs | 6(0.86) | - | - |

## DISCUSSION

The present study is the first surveillance of injury and illness in both athletes and nonathletes participating in Winter Olympic Games, and all patients of the Beijing Winter Olympic Village Polyclinic in the Zhangjiakou Competition Zone and Peking University Third Hospital-Chongli were included^11^. The principal findings were that at least 2.4% of the athletes and nonathletes incurred an injury during the games, and 3.1% of the athletes and nonathletes suffered an illness^12^. The incidences of injuries and illnesses varied substantially between different participant/personnel categories. Athletes tended to sustain acute injuries, while team officials were generally older and more prone to illnesses and degenerative injuries compared with athletes^13^.

### Injuries in the Olympic sports

The aim of the present study was to describe and analyze the athletes’ and nonathletes’ injuries and illnesses that occurred during the Beijing 2022 Winter Olympic Games^14^. The incidence of injury in the Beijing 2022 Winter Olympic Games (2.4%) was lower than that at the Beijing 2008 (10%)^2^, Vancouver 2010 (11%)^3^, London 2012 (11%)^4^, Sochi 2014 (12%)^5^, Rio 2016 (8%)^6^, and PyeongChang 2018 (12%)^7^ Winter Games. The incidence was also lower than the injury incidences reported from the recent Paralympic Games^15-20^. This phenomenon may be owing to the fact that all previous Olympic Games focused on the athletes’ injuries, only. In addition to the athletes’ injuries, we analyzed injuries sustained by nonathletes, in this study. The large sample size reduced the incidence of injuries and illnesses compared with the incidences in previous Winter Olympic Games. The injury rate for athletes at the Beijing Olympics was 14.4% and the illness rate was 4.6%, which were not significantly different from the rates at previous Olympic Games^21^.

### Causes, mechanisms, and circumstances of injury

The causes, mechanisms, and circumstances of injuries sustained during the Beijing Olympic Games differed significantly among different participant/personnel categories. The vast majority of injuries in Beijing were reported as acute, such as contact with stationary objects and sprains, whereas overuse injuries accounted for one-tenth of the injuries. Although similar distributions were previously reported at both Summer and Winter Olympic Games^22^, these numbers should be interpreted with caution owing to limitations in the recording of overuse injuries in the current methodology. In athletes, staff, NTOs and ITOs, and stakeholders, the most commonly reported injury mechanism was contact with a stationary object, while in volunteers and team officials, the most common mechanism of injury was overuse^23^.

### Characteristics of the injury location

Overall, head and neck injuries were the most serious injuries (n=92, 10.7%), and these can be life-threatening. Limb injuries were the most common injuries (n=513, 59.4%), and spinal injuries were the second most common injuries (n=244, 28.2%). In addition to acute sports injuries, degenerative injuries also accounted for a large proportion of the injuries. Injuries to the hips and pelvis were less common (n=34, 4%), probably because some winter sports involve less use of the hips and pelvis. There were some differences regarding the different categories of participants. Lumbar and lower back injuries were common in almost all categories of participants. Additionally, athletes and staff were prone to knee injuries owing to acute injuries or degenerative changes^24^. However, for stakeholders and NTOs and ITOs, the data were not statistically significant owing to the small sample sizes in these groups.

### Analysis of injury types

Fracture (n=169, 19.6%), strain (n=166, 19.2%), and sprain (n=129, 14.9%) were the most common types of injuries in the Beijing Olympic Games. For athletes, volunteers, and team officials, the incidence of strain (muscle rupture, tear, tendon rupture) was highest. However, for staff, the incidence of fracture (trauma, stress, bone contusion, other bone injuries) was highest. These findings suggest that staff need to pay attention to personal protection during their daily work at Olympic venues to avoid unnecessary injuries.

### Characteristics of injury causes

The most common injury causes were contact with a stationary object (n=241, 27.9%) and sprain (n=121, 14.0%). Contact with a stationary object was the most common cause of injury in athletes and staff, while volunteers and team officials were prone to injury from overuse (gradual onset). The injury causes were related to events, occupations, overuse, competition venue, and other factors. Therefore, athletes and nonathletes are recommended to strengthen personal protection measures in the competition venues to avoid unnecessary injuries.

### Characteristics of illness-affected systems, symptoms, and causes

Oral structures and the gastrointestinal system were the most commonly affected systems. The gastrointestinal system was the most commonly affected system in staff, while other categories of participants/personnel were vulnerable to oral injuries. Accordingly, staff were most likely to suffer abdominal pain, while toothache was most common in other categories of personnel. Owing to limitations in the ability to treat some medical conditions in the polyclinic and medical venues, many patients who required tooth extraction or orthodontic treatment could not receive timely and effective treatment. It is suggested that Olympic medical facilities increase their ability to treat oral medical conditions by involving dentists and adding the required medical equipment. Similar to the illness causes reported at previous Olympic Games, infection was the most common cause of illness at the Beijing Winter Olympic Games. Athletes and nonathletes were advised to strengthen their fitness to prevent illness^25^.

### Clinical implications

Our study summarized the injuries and illnesses in athletes and nonathletes at the Beijing 2022 Winter Olympic Games, which is conducive to a comprehensive understanding of the spectrum of injuries and illnesses during the Winter Olympic Games. Our findings indicate the importance of providing more targeted medical service and are conducive to progress in medical care at the Winter Olympic Games. Our findings differ from those in previous studies in that the rate of gastrointestinal and stomatology disorders was higher in nonathletes than that in athletes, in our study. Medical institutions should increase the number of medical personnel in these specialties when providing medical care at the Winter Olympic Games. With the results of this study, we hoped to provide a reference for the formulation of diagnostic and treatment strategies for injuries and illnesses in both athletes and nonathletes participating in the Winter Olympic Games.

### Limitations and challenges

Our data were derived from all patients who visited the polyclinic and medical venues during the Beijing 2022 Winter Olympics. This method provides a comprehensive overview of the medical encounters of both athletes and nonathletes. However, it would be ideal to obtain follow-up results in future studies.

## CONCLUSION

The injury morbidity rate of athletes at the Beijing 2022 Winter Olympics was 14.4%, and the illness morbidity rate was 4.6%. For nonathletes, the injury morbidity rate was 2.0%, and the illness morbidity rate was 3.1%. The incidence of injuries and illnesses varied substantially between the different participant/personnel categories. Analyses of injury mechanisms and illness causes for both athletes and nonathletes during the Winter Olympic Games are essential to devise better injury and illness prevention strategies. This study provides a basis for the formulation of these strategies for future Winter Olympic Games.

## Data Availability

All data relevant to the study are included in the article.

## Acknowledgements

We would like to acknowledge the contribution and support of the Beijing 2022 Winter Olympic Games staff throughout the different stages of this study, particularly the medical team and Beijing Organizing Committee for the 2022 Olympic and Paralympic Winter Games. Additionally, we sincerely thank all the medical staff who contributed to the data collection.

## Contributors

All authors contributed to the study conception and design, and data collection and interpretation. Zhen-long Liu and Can-can Du analyzed the data and drafted the paper. All authors provided revisions and contributed to the final manuscript. Zhen-long Liu and Can-can Du are the guarantors.

## Patient and public involvement

Patients and the public were not involved in the design or conduct of this research.

## Patient consent for publication

Not required.

## Ethics approval

This study was reviewed and approved by the Peking University Third Hospital Medical Science Research Ethics Committee (M2022428).

## Provenance and peer review

Not commissioned; externally peer reviewed.

## Data availability statement

All data relevant to the study are included in the article.

## REFERENCE

1. Junge A, Engebretsen L, Alonso JM, et al. Injury surveillance in multi-sport events: the International Olympic Committee approach. Br J Sports Med 2008; 42:413–21

2. Junge A, Engebretsen L, Mountjoy ML, et al. Sports injuries during the Summer Olympic Games 2008. Am J Sports Med 2009; 37:2165–72

3. Engebretsen L, Steffen K, Alonso JM, et al. Sports injuries and illnesses during the Winter Olympic Games 2010. Br J Sports Med 2010; 44:772–80

4. Engebretsen L, Soligard T, Steffen K, et al. Sports injuries and illnesses during the London Summer Olympic Games 2012. Br J Sports Med 2013; 47:407–14

5. Soligard T, Steffen K, Palmer-Green D, et al. Sports injuries and illnesses in the Sochi 2014 Olympic Winter Games. Br J Sports Med 2015; 49:441–7

6. Soligard T, Steffen K, Palmer D, et al. Sports injury and illness incidence in the Rio de Janeiro 2016 Olympic Summer Games: A prospective study of 11274 athletes from 207 countries. Br J Sports Med 2017; 51:1265–71

7. Soligard T, Palmer D, Steffen K, et al. Sports injury and illness incidence in the PyeongChang 2018 Olympic Winter Games: a prospective study of 2914 athletes from 92 countries. Br J Sports Med 2019; 53:1085–92

8. Bull FC, Al-Ansari SS, Biddle S, et al. World Health Organization 2020 guidelines on physical activity and sedentary behaviour. Br J Sports Med 2020; 54:1451–62

9. Mountjoy M, Junge A, Alonso JM, et al. Consensus statement on the methodology of injury and illness surveillance in FINA (aquatic sports). Br J Sports Med 2016; 50:590–6

10. Clarsen B, Bahr R. Matching the choice of injury/illness definition to study setting, purpose and design: one size does not fit all! Br J Sports Med 2014; 48:510–2

11. Zwiers R, Zantvoord FW, Engelaer FM, et al. Mortality in former Olympic athletes: retrospective cohort analysis. BMJ 2012; 345:e7456

12. Fuller CW, Taylor A, Raftery M. 2016 Rio Olympics: an epidemiological study of the men’s and women’s Rugby-7s tournaments. Br J Sports Med 2017; 51:1272–78

13. Clarke PM, Walter SJ, Hayen A, et al. Survival of the fittest: retrospective cohort study of the longevity of Olympic medallists in the modern era. Br J Sports Med 2015; 49:898–902

14. S Drawer CWF. Evaluating the level of injury in English professional football using a risk based assessment process. Br J Sports Med 2002; 36:446–51

15. Derman W, Runciman P, Schwellnus M, et al. High precompetition injury rate dominates the injury profile at the Rio 2016 Summer Paralympic Games: a prospective cohort study of 51 198 athlete days. Br J Sports Med 2018; 52:24–31

16. Derman W, Schwellnus M, Jordaan E, et al. Illness and injury in athletes during the competition period at the London 2012 Paralympic Games: development and implementation of a web-based surveillance system (WEB-IISS) for team medical staff. Br J Sports Med 2013; 47:420–5

17. Derman W, Schwellnus MP, Jordaan E, et al. The incidence and patterns of illness at the Sochi 2014 Winter Paralympic Games: a prospective cohort study of 6564 athlete days. Br J Sports Med 2016; 50:1064–8

18. Webborn N, Cushman D, Blauwet CA, et al. The Epidemiology of Injuries in Football at the London 2012 Paralympic Games. PM R 2016; 8:545–52

19. Webborn N, Willick S, Emery CA. The Injury Experience at the 2010 Winter Paralympic Games. Clin J Sport Med 2012; 22:3–9

20. Webborn N, Willick S, Reeser JC. Injuries among disabled athletes during the 2002 Winter Paralympic Games. Med Sci Sports Exerc 2006; 38:811–5

21. Clarsen B, Ronsen O, Myklebust G, et al. The Oslo Sports Trauma Research Center questionnaire on health problems: a new approach to prospective monitoring of illness and injury in elite athletes. Br J Sports Med 2014; 48:754–60

22. Teramoto M, Bungum TJ. Mortality and longevity of elite athletes. J Sci Med Sport 2010; 13:410–16

23. Clarsen B, Myklebust G, Bahr R. Development and validation of a new method for the registration of overuse injuries in sports injury epidemiology: the Oslo Sports Trauma Research Centre (OSTRC) overuse injury questionnaire. Br J Sports Med 2013; 47:495–502

24. S Drawer CWF. Propensity for osteoarthritis and lower limb joint pain in retired professional soccer players. Br J Sports Med 2001; 35:402–08

25. Bahr R. No injuries, but plenty of pain? On the methodology for recording overuse symptoms in sports. Br J Sports Med 2009; 43:966–72

